# Rare Missense Variants in *MYO7A* and *OTOP2* Genes in a South Korean Meniere Disease Cohort

**DOI:** 10.1101/2025.06.16.25329383

**Authors:** Mai T. Pham, Pablo Cruz-Granados, Seung Hyun Jang, Heon Yung Gee, Jinsei Jung, Jae Young Choi, Sung Huhn Kim, Jose A. Lopez-Escamez

## Abstract

Meniere disease (MD) is a polygenic condition defined by episodes of vertigo associated with sensorineural hearing loss and tinnitus. Genetic studies in familial MD in East Asian population are limited and the potential MD genes remain to be established in non-Finnish European populations. By exome sequencing and rare variant analysis, we search for existing and novel genes associated with MD in a South Korean cohort of 16 MD individuals with bilateral sensorineural hearing loss. We have found one individual with two rare missense variants in the *OTOP2* gene, a new candidate gene for MD and three heterozygous variants in the *MYO7A* gene, supporting the hypothesis of biallelic inheritance. Protein modelling was conducted on three rare missense variants in *OTOP2* to further elucidate functional consequences. The structural and functional implications inferred from these models suggest a likely pathogenic role, providing additional insights into the molecular mechanisms underlying MD.

## INTRODUCTION

Meniere’s disease (MD) is a rare inner ear disorder, typically characterised by sensorineural hearing loss (SNHL), episodic vertigo, tinnitus, and aural fullness [1]. It has a significant heritability according to epidemiological and genetic studies [2,3]. Exome sequencing studies have identified approximately 20 familial MD genes, most of which are also associated with syndromic and non-syndromic SNHL, notably *OTOG, MYO7A*, and *TECTA* [4–8].

Familial MD has a complex inheritance with autosomal dominant mediated by one loss-of-function (LoF) variant with incomplete penetrance [9], or autosomal recessive, including digenic inheritance mediated by two or more missense variants [5,6]. The most common gene found is *OTOG* which encodes otogelin, an extracellular protein involved in anchoring hair cell stereocilia to the tectorial membrane in the organ of Corti [4]. *OTOP2* has recently emerged as a MD candidate gene, tightly linked to vestibular dysfunction [10]. A rare missense variant in *OTOP2* was also reported in a Spanish MD proband [10].

Most genetic studies on MD have been conducted in European population, particularly in Spain, and the genetic contribution in Asian population has primarily been assessed using gene panels [11]. Therefore, replication studies of MD cases and its related clinical characteristics in Asian population, particularly in East Asian (EAS), are necessary to elucidate shared genes and inheritance patterns of MD across different populations.

Here, we present exome sequencing data from a South Korean MD cohort (SK-MD, n = 16). By comparing allele frequencies with the EAS population of the gnomAD database, we have identified a new candidate gene, *OTOP2*, with a burden of rare missense variants. Notably, two missense variants in *OTOP2* gene have been found in the same MD individual. This study provides support for the heterozygous compound recessive and digenic inheritance patterns in MD, further refining our understanding of its genetic architecture.

## MATERIAL AND METHODS

### Patient Selection

Participants were selected according to the 2015 Barany Society diagnostic criteria for MD [12]. The familial history of SNHL and episodic vertigo was obtained to generate pedigrees.

### DNA Extraction and exome sequencing

Exome sequencing was conducted on 16 SK-MD samples (from Yonsei University Hearing Loss cohort) obtained from blood or saliva, using the Easy-DNA™ Kit (Invitrogen Corporation, Carlsbad, CA, USA) according to the manufacturer’s protocol and quality control (QC) procedures, as previously described [13,14]. DNA libraries were subjected to exome capture using a SureSelect V5 enrichment capture kit (Agilent Technologies, Santa Clara, CA, USA). The enriched library was then sequenced on an HiSeq 2500 platform (101 bases, paired-end) or NovaSeq 6000 sequencing platform (151 bases, paired-end) (Illumina, San Diego, CA, USA) as described previously [15].

### Bioinformatic Analysis

The pipeline consisted of two main steps: preprocessing and variant calling for germline single nucleotide variants (SNVs) and short indels, following the GATK best practice (v4.6.1.0) [16]. A summary of workflow is presented in Supplementary Figure S1.

### Quality control and read alignment

Raw RNA-seq reads were evaluated for QC using *FASTQC* (v0.12.1) and pre-processed with *Cutadapt* (v1.18) [17] to remove Illumina adapter sequences and low-quality bases. The cleaned reads were then aligned to the human reference genome hg38 (GATK resource bundle) using *bwa-mem2* (v2.2.1) [18]. Aligned reads in BAM files were then marked duplicates and Base Quality Score Recalibration (BQSR) using *picards MarkDuplicates* [19] and *gatk BaseCalibrator/applyBQSR*, respectively. Known polymorphic sites from the GATK resource bundle were used to build recalibration tables, subsequently correcting for systematic biases in base quality scores introduced by the sequencing process.

### Germline SNV and indel calling and filtering

*Gatk HaplotypeCaller* was used for per-sample variant calling, followed by *CombineGVCF/GenotypeGVCF* for joint calling with genotypic information for each sample. Variant filtering was then applied using *gatk VariantCalibrator/applyVQSR* to obtain high-quality variant candidates with a sensitivity threshold of 90% for all SNV and indels, based on known SNV/indel databases and machine learning modelling. As recommended in GATK VQSR strategy, SNV VQSR model was trained using single-nucleotide polymorphism (SNP) sites from HapMap (v3.3), high-confidence SNPs in 1000 Genome Project (1000GP), Illumina Omni 2.5M SNP arrays, and dbSNP (v138). The indel VQSR model was trained using high-confidence indel sites from 1000GP and dbSNP (v138). Any variants marked as ‘PASS’ from VQSR filtering were subsequently filtered using hard-filter parameters, as recommended by GATK best practice to remove low-confident variants. We used *gatk VariantFiltration* to filter SNVs based on the following criteria: Quality by Depth (QD) < 2.0, Strand Odds Ratio (SOR) > 3.0, variant QUAL < 30, Fisher Strand (FS) > 60, Mapping Quality (MQ) < 40.0, Mapping Quality Rank Sum Test (MQRankSum) < -12.5, and Read Position Rank Sum Test (ReadPosRankSum) < -6.0. Indels were filtered using QD < 2.0, FS > 200, QUAL < 30, and ReadPosRankSum < -20. Passed variants were further filtered to retain rare variants with an allele frequency (AF) < 0.05 for downstream annotation and statistical analyses.

### Variant annotation and pathogenicity assessment

The *Predictor VEP* tool (v113) [20] was used for variant annotations. We categorised exonic SNVs based on their protein consequences, including missense, loss-of-function (LoF) (including stop gained, stop/start loss, splice donor/acceptor). For short indels, we only considered frameshift mutations.

We then selected variants located in known and potential MD genes [6,10]. We then used the Combined Annotation Dependent Depletion (CADD) database [21] to retrieve CADD Phred-scaled score. Selected variants were interpreted using both the 2015 American College Medical Genetics/Association for Molecular Pathology (ACMG/AMP) guidelines (via the GeneBe database [22]) and the Hearing Loss Expert Panel (HL-EP)-specific guidelines (from the ClinGen Evidence Repository [23]).

AlphaMissese [24] pathogenicity predictions were used to generate heatmaps. For selected genes, a sliding window consisting of 200 base-pair (bp) upstream and downstream of the chosen mutations were created to identify constraint regions with an overload of missense variants. GnomAD v2.1 [25] was used to retrieve missense variants in the genes of interest for each population (EAS, South Asian – SAS, Finnish European – FIN, Non-Finnish European – NFE, Ashkenazi Jew – ASH, Sub-Sharan African – SSA, and Admixed American – AMR). To assess pathogenicity of the selected mutations, expected number of missense variants per population were retrieved from gnomAD v2.1, and were used to estimate a high-density threshold.

*SpliceAI* [26] and *Pangolin* [27] were employed to predict splice sites within the selected proteins. A 300 bp window was considered upstream and downstream of the variant positions. Additionally, *Human Splice Finder Pro* (HSF Pro) was used to predict DNA sequence motifs associated with splicing [28].

### Statistical Analysis

We performed a gene burden analysis (GBA) to search for genes with a burden of rare exonic variants associated with SK-MD cohort. These variants are rare missense and LoF SNVs, as well as frameshift indels that possess moderate-to-high effects in functional consequences.

We retrieved allele counts, allele number, and AF of ‘PASS’ variants from the exome data of gnomAD database v4.1 for the EAS (EAS-gnomAD, n = 19,850) and global populations (global-gnomAD, n = 730,947), served as control groups for GBA. For each gene, aggregated AFs, calculated based on the total number of alternate alleles, were compared between the SK-MD cohort (case group, n = 16) and each control group using Fisher exact’s test to assess gene-level burden associations. Odd ratio (OR) with a 95% confidence interval (CI), aetiology fraction (EF), and two-sided *P*-value were recorded. Adjusted *P*-values using Bonferroni correction were obtained to account for total genes. Variants with adjusted *P* < 0.05 and *OR* > 1 were significantly enriched in a specific population. Frequently mutated genes (FLAGS) were then excluded from further downstream analyses [29].

For biallelic digenic mutation, we selected genes encoding proteins with physical interaction and calculated combined minor AF (cMAF) for EAS-gnomAD, global-gnomAD, and our SK-MD cohort by multiplying two MAF values of two missense variants, with the assumption of independency of both variants. Association testing between our SK-MD cohort and gnomAD control group using Fisher’s exact test with OR with a 95% CI and two-sided *P*-value after Bonferroni correction was also recorded.

### Protein Modelling

PBD files for wild type (wt) proteins were retrieved from AlphaFold2 Protein Structure Database [30] and were assessed for high quality. Sequences were retrieved from UniProt Database [31] to generate mutant (mt) protein models by homology using *MODELLER* v10.6 [32]. Mutant protein models were selected based on the built-in *MODELLER* quality controls; DOPE and GA341 scores.

Quality control checks were performed atomic interactions and stereochemical quality in both the wt and mt proteins using *ERRAT* [33], *WHATCHECK* (https://swift.cmbi.umcn.nl/gv/whatcheck/index.html) and *PROCHECK* [34] from the Saves Server v.6.1 (https://saves.mbi.ucla.edu/).

Protein stability and atomic interactions were predicted using *DinaMut2* [35], and model visualisation was done employing *PyMOL* Open Source (*Schrodinger*, LLC. 2010. The PyMOL Molecular Graphics System, Version 3.0.x.).

## RESULTS

### Burden of rare variants in South Korean individuals with MD

By comparing the SK-MD cohort with the reference datasets from the EAS and global gnomAD populations, we found that most significant gene burdens were driven by rare missense variants (AF < 0.05). Gene burden observed when comparing with reference AF from non-EAS populations was not considered EAS population-specific and were therefore excluded from further analysis.

We found 14 genes with a significant gene burden of rare variants associated with MD across different populations (Table S1). The top four genes showing the strongest gene burden in the EAS population (adjusted *P* < 0.01) were *CCNJL* (*OR* = 774.41 [117.33 – 3884.09], adjusted *P* = 1.30 x 10^-4^), *BAHCC1* (*OR* = 71.61 [18.6 – 196.47], adjusted *P* = 2.26 x 10^-3^), *IGFN1* (*OR* = 19.7 [7.09 – 44.14], adjusted *P* = 4.58 x 10^-3^), and *CDON* (*OR* = 2436.47 [130.24 – 4.5 x 10^15^], adjusted *P* = 8.80 x 10^-3^). Among these, *BAHCC1* showed a significant population-specific burden in the EAS population. Of note, *IGFN1*, *CDON*, and *OTOP2* (*OR* = 100.88 [19.6 – 326.87], adjusted *P* = 2.43 x 10^-2^) each harboured two rare missense variants in the same individual, suggesting potential compound heterozygous inheritance. However, no single variant was significantly associated with MD in the EAS population.

### Heterozygous Missense Variants in MD genes

We found one SK-MD individual, P1, carrying two heterozygous missense variants (chr17:74930726:C>T and chr17:74930940:C>G) in *OTOP2*, indicating a compound recessive inheritance pattern (Table 1). Besides, a rare missense variant (chr11:77156886:G>A, CADD = 26.6) in *MYO7A* was identified in individual P2, co-occurring with a second missense variant (chr10:71778252:G>A, CADD = 20.8) in *CDH23*, suggesting potential digenic inheritance in the SK-MD cohort (Table 2). According to the 2015 ACMG/AMP and the 2018 HL-EP guidelines, the variant chr10:71778252:G>A in *CDH23* was classified as likely benign and it has been implicated in Usher syndrome with an autosomal recessive inheritance pattern (ClinVar ID: 197422). Individual P2 also carried a third rare missense variant (chr17:74930450:C>T) in *OTOP2*.

**Table 1:**
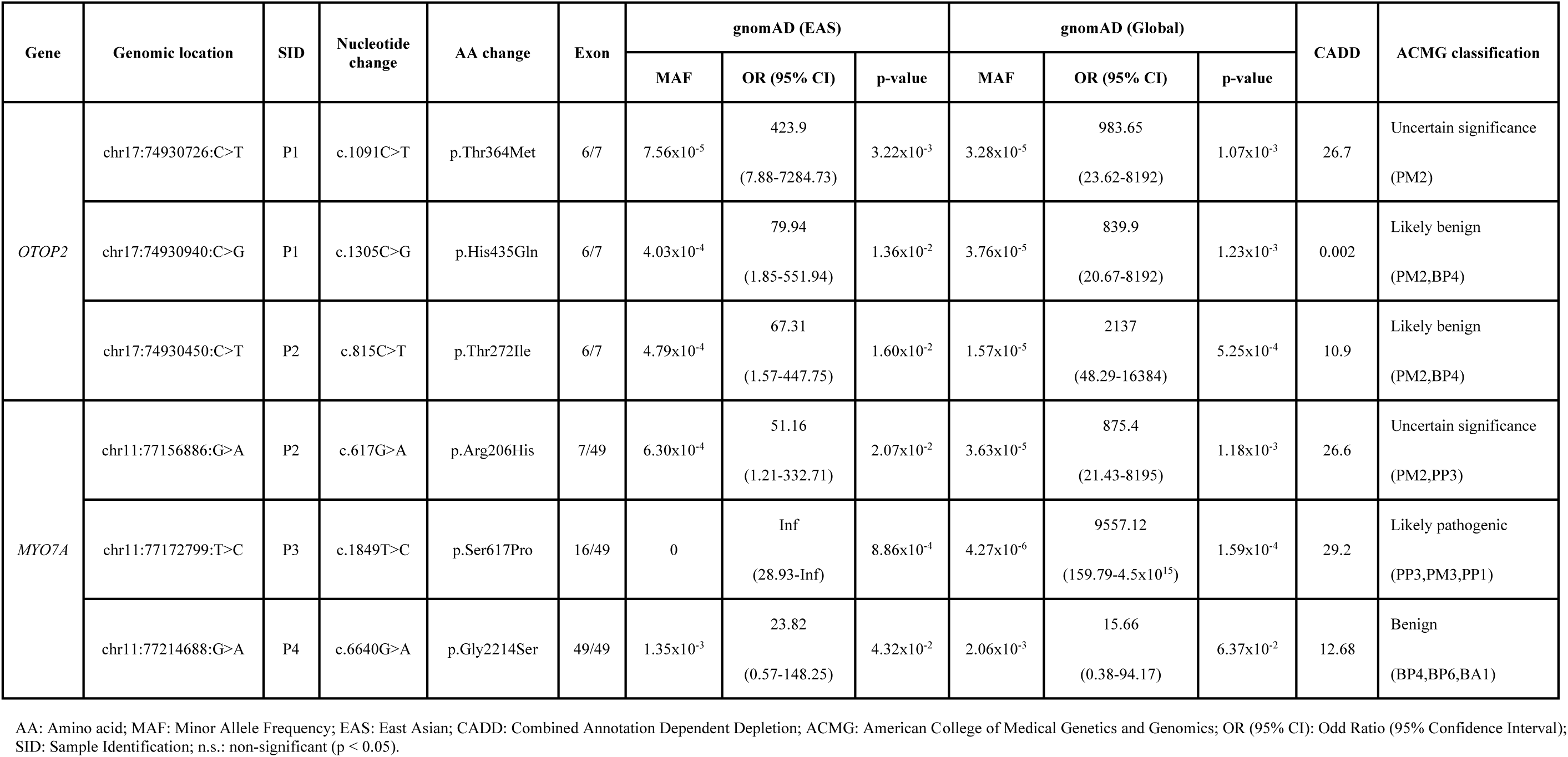
Missense rare variants found in the *MYO7A* and *OTOP2* genes in the MD-SK cohort. Biallelic mutations in *OTOP2* gene observed in a single SK-MD individual (P1).

| Gene | Genomic location | SID | Nucleotide change | AA change | Exon | gnomAD (EAS) |  |  | gnomAD (Global) |  |  | CADD | ACMG classification |
| --- | --- | --- | --- | --- | --- | --- | --- | --- | --- | --- | --- | --- | --- |
|  |  |  |  |  |  | MAF | OR (95% CI) | p-value | MAF | OR (95% CI) | p-value |  |  |
| *OTOP2* | chr17:74930726:C>T | P1 | c.1091C>T | p.Thr364Met | 6/7 | 7.56x10<sup>-5</sup> | 423.9<br>(7.88-7284.73) | 3.22x10<sup>-3</sup> | 3.28x10<sup>-5</sup> | 983.65<br>(23.62-8192) | 1.07x10<sup>-3</sup> | 26.7 | Uncertain significance<br>(PM2) |
|  | chr17:74930940:C>G | P1 | c.1305C>G | p.His435Gln | 6/7 | 4.03x10<sup>-4</sup> | 79.94<br>(1.85-551.94) | 1.36x10<sup>-2</sup> | 3.76x10<sup>-5</sup> | 839.9<br>(20.67-8192) | 1.23x10<sup>-3</sup> | 0.002 | Likely benign<br>(PM2,BP4) |
|  | chr17:74930450:C>T | P2 | c.815C>T | p.Thr272Ile | 6/7 | 4.79x10<sup>-4</sup> | 67.31<br>(1.57-447.75) | 1.60x10<sup>-2</sup> | 1.57x10<sup>-5</sup> | 2137<br>(48.29-16384) | 5.25x10<sup>-4</sup> | 10.9 | Likely benign<br>(PM2,BP4) |
| *MYO7A* | chr11:77156886:G>A | P2 | c.617G>A | p.Arg206His | 7/49 | 6.30x10<sup>-4</sup> | 51.16<br>(1.21-332.71) | 2.07x10<sup>-2</sup> | 3.63x10<sup>-5</sup> | 875.4<br>(21.43-8195) | 1.18x10<sup>-3</sup> | 26.6 | Uncertain significance<br>(PM2,PP3) |
|  | chr11:77172799:T>C | P3 | c.1849T>C | p.Ser617Pro | 16/49 | 0 | Inf<br>(28.93-Inf) | 8.86x10<sup>-4</sup> | 4.27x10<sup>-6</sup> | 9557.12<br>(159.79-4.5x10<sup>15</sup>) | 1.59x10<sup>-4</sup> | 29.2 | Likely pathogenic<br>(PP3,PM3,PP1) |
|  | chr11:77214688:G>A | P4 | c.6640G>A | p.Gly2214Ser | 49/49 | 1.35x10<sup>-3</sup> | 23.82<br>(0.57-148.25) | 4.32x10<sup>-2</sup> | 2.06x10<sup>-3</sup> | 15.66<br>(0.38-94.17) | 6.37x10<sup>-2</sup> | 12.68 | Benign<br>(BP4,BP6,BA1) |

AA: Amino acid; MAF: Minor Allele Frequency; EAS: East Asian; CADD: Combined Annotation Dependent Depletion; ACMG: American College of Medical Genetics and Genomics; OR (95% CI): Odd Ratio (95% Confidence Interval); SID: Sample Identification; n.s.: non-significant (p < 0.05).

**Table 2:** Individual and combined allelic frequencies of rare variants in the *MYO7A, CDH23 and ADGRV1* genes encoding proteins involved in the structure of hair cell stereocilia links. Minor Allelic Frequencies (MAF) and odds ratio (OR) were calculated using reference data from the EAS and global populations in the gnomAD database.

|  | Genomic location | SID | cMAF | gnomAD (Global) |  |  |  | gnomAD (EAS) |  |  |  | CADD | ACMG classification |
| --- | --- | --- | --- | --- | --- | --- | --- | --- | --- | --- | --- | --- | --- |
|  |  |  |  | MAF | cMAF | OR (95% CI) | adj. P-value | MAF | cMAF | OR (95% CI) | adj. P-value |  |  |
| MYO7A | chr11:77156886:G>A | P2 | 9.77x10<sup>-4</sup> | 3.63x10<sup>-5</sup> | 5.74x10<sup>-9</sup> | 16383.63<br>(560.98-4.50x10<sup>15</sup>) | 9.04x10<sup>-5</sup> | 6.30x10<sup>-4</sup> | 1.19x10<sup>-6</sup> | 1212.627<br>(15.33-4.50x10<sup>15</sup>) | 3.32x10<sup>-3</sup> | 26.6 | Uncertain significance |
| CDH23 | chr10:71778252:G>A |  |  | 1.58x10<sup>-4</sup> |  |  |  | 1.89x10<sup>-3</sup> |  |  |  | 20.8 | Likely benign |
| MYO7A | chr11:77214688:G>A | P4 | 9.77x10<sup>-4</sup> | 2.06x10<sup>-3</sup> | 3.52x10<sup>-8</sup> | 16383.62<br>(548.09- 4.50x10<sup>15</sup>) | 9.28x10<sup>-5</sup> | 1.35x10<sup>-3</sup> | 8.51x10<sup>-7</sup> | 1129.14<br>(14.57-4.50x10<sup>15</sup>) | 3.50x10<sup>-3</sup> | 12.68 | Benign |
| ADGRV1 | chr5:90672678:A>G |  |  | 1.71x10<sup>-5</sup> |  |  |  | 6.30x10<sup>-4</sup> |  |  |  | 24.4 | Likely benign |

MAF: Minor Allele Frequency; cMAF: combined MAF; EAS: East Asian; OR (95% CI): Odd Ratio (95% Confidence; adj. P-value: Adjusted P-value after Bonferroni correction; CADD: Combined Annotation Dependent Depletion; ACMG: American College of Medical Genetics and Genomics; Interval); SID: Sample Identification

The clinical profiles of two MD patients carrying rare variants in these genes are described below (Figure 1). Full pedigrees not shown in Figure 1 to avoid potential identification of the families.

- P1 was diagnosed with MD in their mid-twenties, following an adult onset at their early twenties with hearing loss and dizziness, accompanied with migraine, diabetes insipidus, fibromyalgia, and depression. They received an endolymphatic sac surgery after five years since the onset of symptoms. They were the only individual with MD in the multi-generational family, which includes a few members with SNHL and/or tinnitus. The individual carried two heterozygous missense variants in *OTOP2*.
- P2 had no other relatives showing MD symptoms, although one of their descendants suffered tinnitus. They were diagnosed with MD in their fifties, following a late onset with symptoms that started one year before (dizziness and progressive bilateral hearing loss, along with comorbid hypertension). They received a cochlear implant in the right ear after diagnosis. The individual exhibited a missense variant in *OTOP2*, as well as rare missense variants in the genes *MYO7A* and *CDH23*.

**Figure 1:**
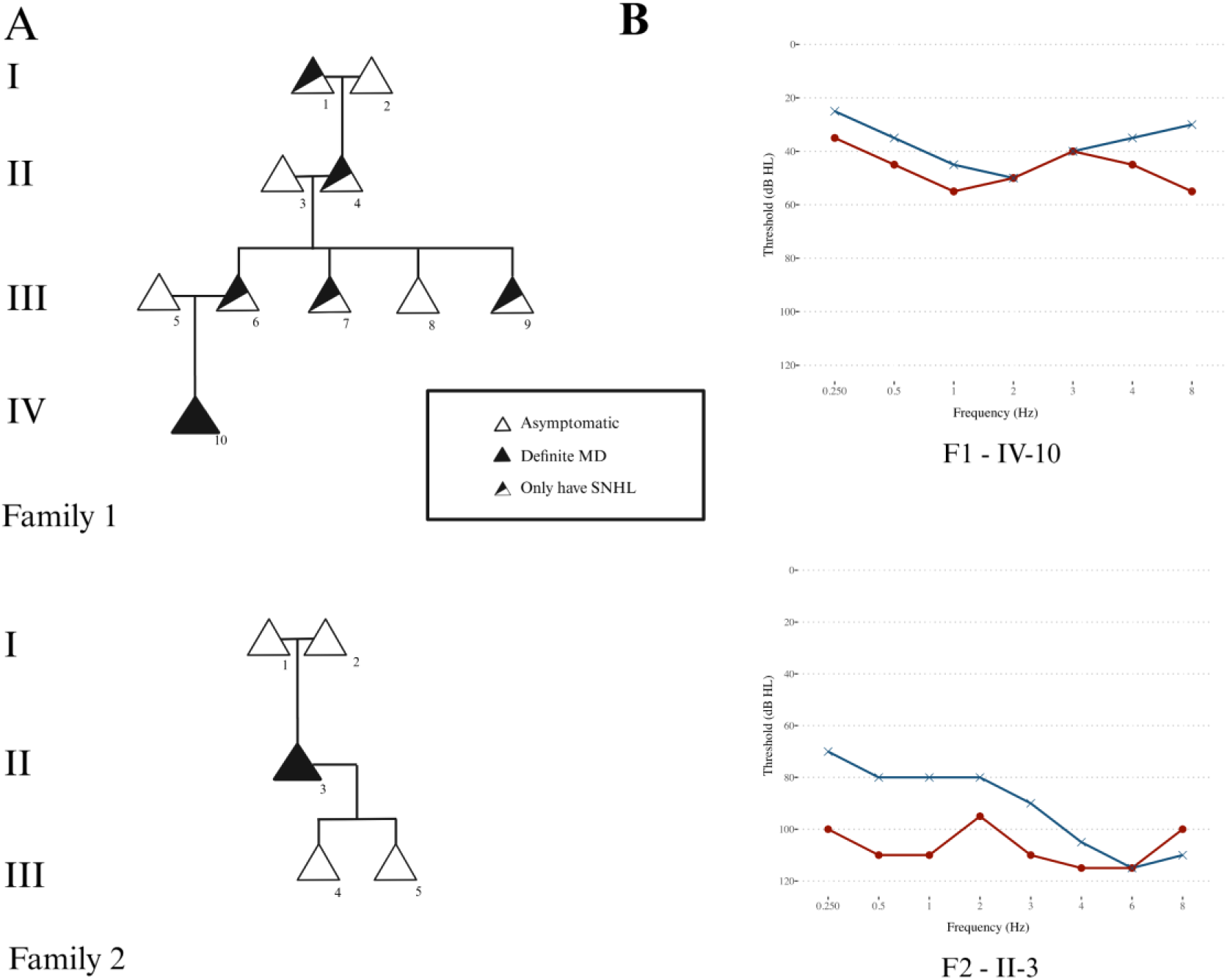
Pedigrees of two patients carrying rare missense variants in *OTOP2*: Family 1 (Individual P1 – F1-IV10), Family 2 (Individual P2 – F2-II3). SNHL: Sensorineural hearing loss.

In addition, another two SK-MD presented missense variants in *MYO7A*. P3 harboured a rare missense variant in *MYO7A* (chr11:77172799:T>C), which has not been previously reported in the EAS population and is exceptionally rare globally (MAF = 4.27 x 10^-6^). According to the 2015 ACMG/AMP and the 2018 HL-EP guidelines, this variant is likely pathogenic (CADD score = 29.2) and has been implicated in autosomal recessive non-syndromic deafness (ClinVar ID: 438172). A four individual (P4) with another rare missense variant in *MYO7A* (chr11:77214688:G>A, CADD = 12.68) also had a second missense variant (chr5:90672678:A>G, CADD = 24.4) in the gene *ADGRV1*. The occurrence of both variants in the SK-MD was significantly high compared with the expected MAF in the EAS population (cMAF_SK-MD_ = 9.77 x 10^-4^ , cMAF_EAS-gnomAD_ = 8.51 x 10^-7^, *OR* = 1129.14 [14.57 – 4.50 x 10^15^], adjusted *P* = 3.50 x 10^-3^) (Table 2).

### Missense Variants in *OTOP2*

*OTOP2* was the only gene found with rare biallelic missense variants in the same individual (P1) in the SK-MD cohort. Three rare missense variants of *OTOP2* were identified in two unrelated SK-MD individuals. Two heterozygous variants, chr17:74930726:C>T (p.Thr364Met) as a variant of uncertain significance and chr17:74930940:C>G (p.His435Gln), were found in the individual P1, while another heterozygous variant, chr17:74930450:C>T (p.Thr272Ile), was found in the individual P2.

Heatmaps created from AlphaMissese pathogenicity predictions showed the three missense variants as likely benign, with chr17:74930450:C>T having a score of 0.088, chr17:74930726:C>T a score of 0.141, and chr17:74930940:C>G a score of 0.079 (Supplementary Figs. S2-S4). Pathogenicity assessment employing a sliding window of constraint regions revealed that all three variants were in high-density regions in the EAS population (Figure 2A). However, mutations chr17:74930450:C>T and chr17:74930940:C>G were found in border line high-density regions in EAS, very close to low-density gaps in the coding sequence. The three variants were located in both low and high-density regions across the remaining populations.

**Figure 2:**
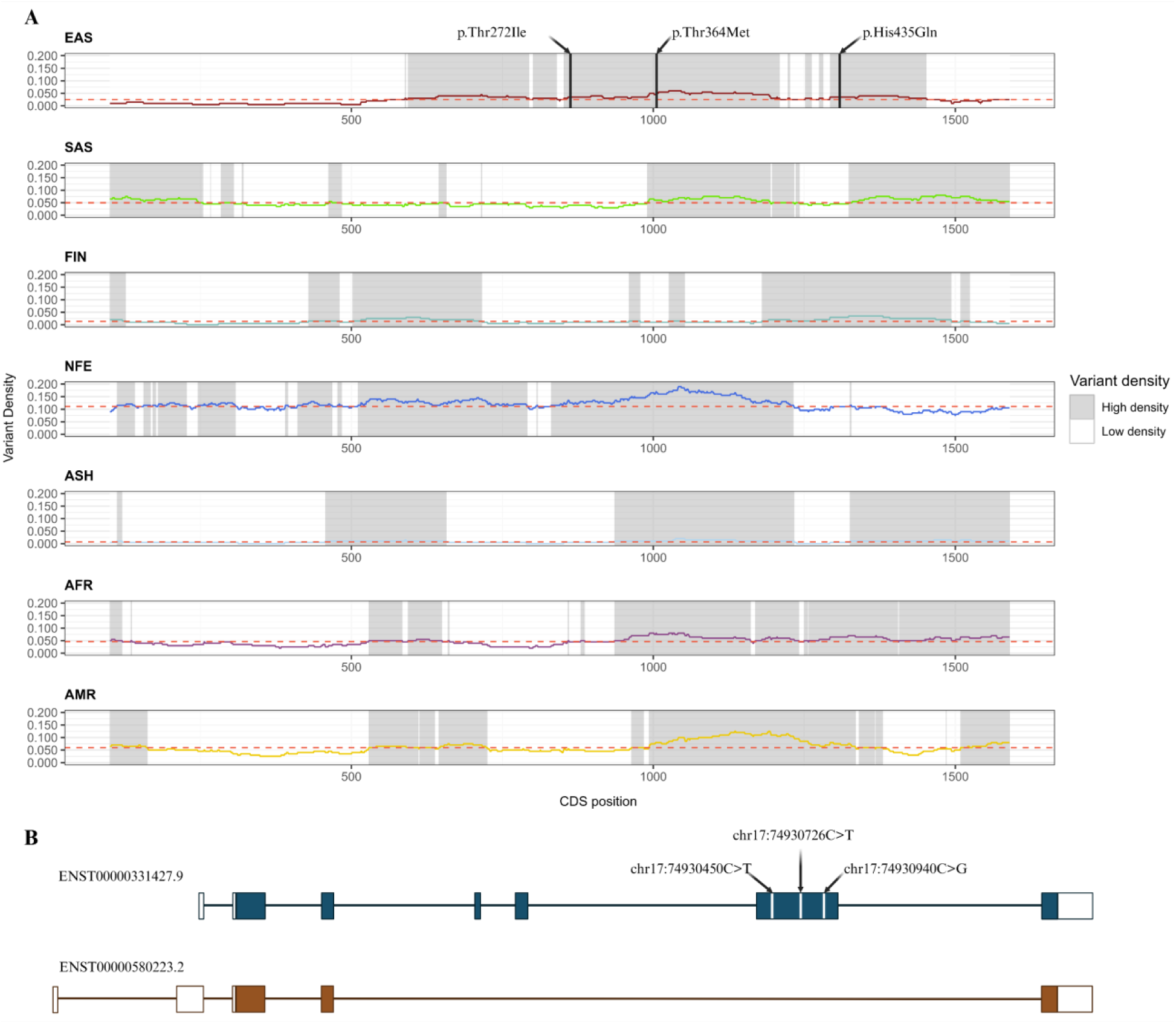
Structural overview of human *OTOP2* gene with missense variants. (A) Low/High density regions across different control gnomAD subpopulations; (B) Locations of three missense variants in the exon 6 of the MANE transcript (ENST00000331427.9).

*OTOP2* has two protein-coding transcripts according to Ensembl (https://ensembl.org). The three variants found in the SK-MD cohort are in exon 6/7 in the MANE transcript (ENST00000331427.9), but not in the alternative transcript ENST00000580223.2 (Figure 2B). While *SpliceAI* and *Pangolin* predicted no splice sites for neither of three variants on a 300-bp window (Table S2), *HSF Pro* predicted a new acceptor splice site starting in chr17:74930929 and finishing in chr17:74930942, resulting from *OTOP2* chr17:74930940:C>G mutation (Table 3). Furthermore, creation and breakage of exonic splice enhancers (ESE) and silencers (ESS) were predicted for all three variants (Table S3).

**Table 3:** Splicing prediction of the variant chr17:74930940:C>G of *OTOP2*, according to *Human Splice Finder Pro* (HSF Pro).

| Human Splicing Finder Matrix |  |
| --- | --- |
| Position | chr17:74930929 |
| Sequences | GCTGAGCCTCACAG>GCTGAGCCTCAGAG |
| Variation | 48.32>76.19 (57.68%) |
| Signal | New Acceptor splice site |
| Interpretation | Activation of a cryptic Acceptor site. Potential alteration of splicing |
| Associated transcripts | ENST00000331427;ENST00000580223 |

### *OTOP2* Protein Models

*OTOP2* canonical transcript ENST00000331427.9 was chosen to model the protein, since it includes exon 6, where the three mutations are located. Modelling of Proton Channel OTOP2 protein (*OTOP2* gene) revealed that amino acid change of p.Thr272Ile (chr17:74930450:C>T) (Figure 3A) and p.His435Gln (chr17:74930726:C>T) were in a loop region, away from the main alpha helices (Figure 4A). On the other hand, the variant p.Thr364Met (chr17:74930940:C>G) was located at the end of one of the alpha helix chains.

**Figure 3:**
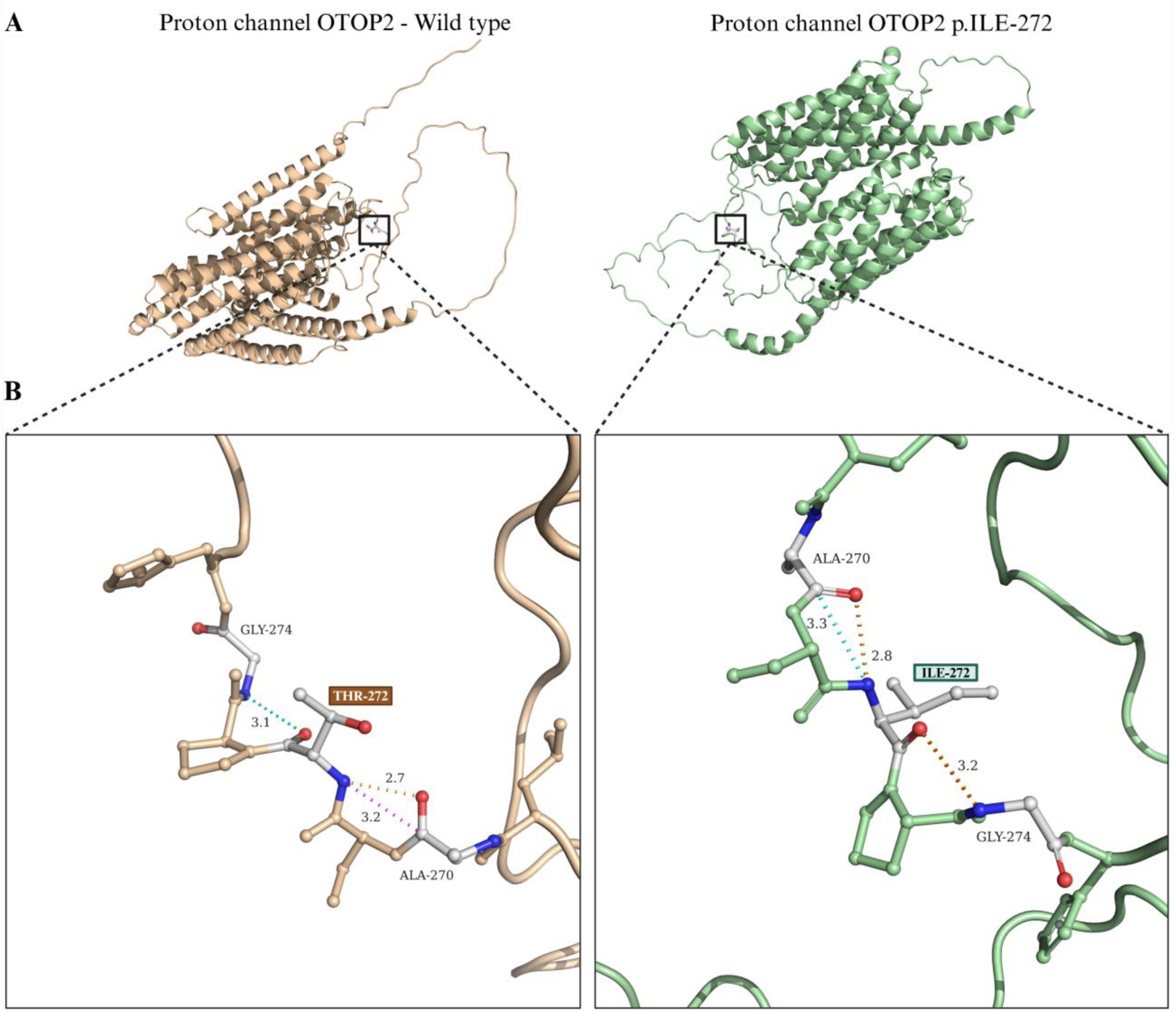
(A) Wild type (beige) and mutant (green) proton channel OTOP2 protein models. (B) Atomic interactions between wild type Thr272 and mutant Ile272. Mutated amino acids are highlighted.

**Figure 4:**
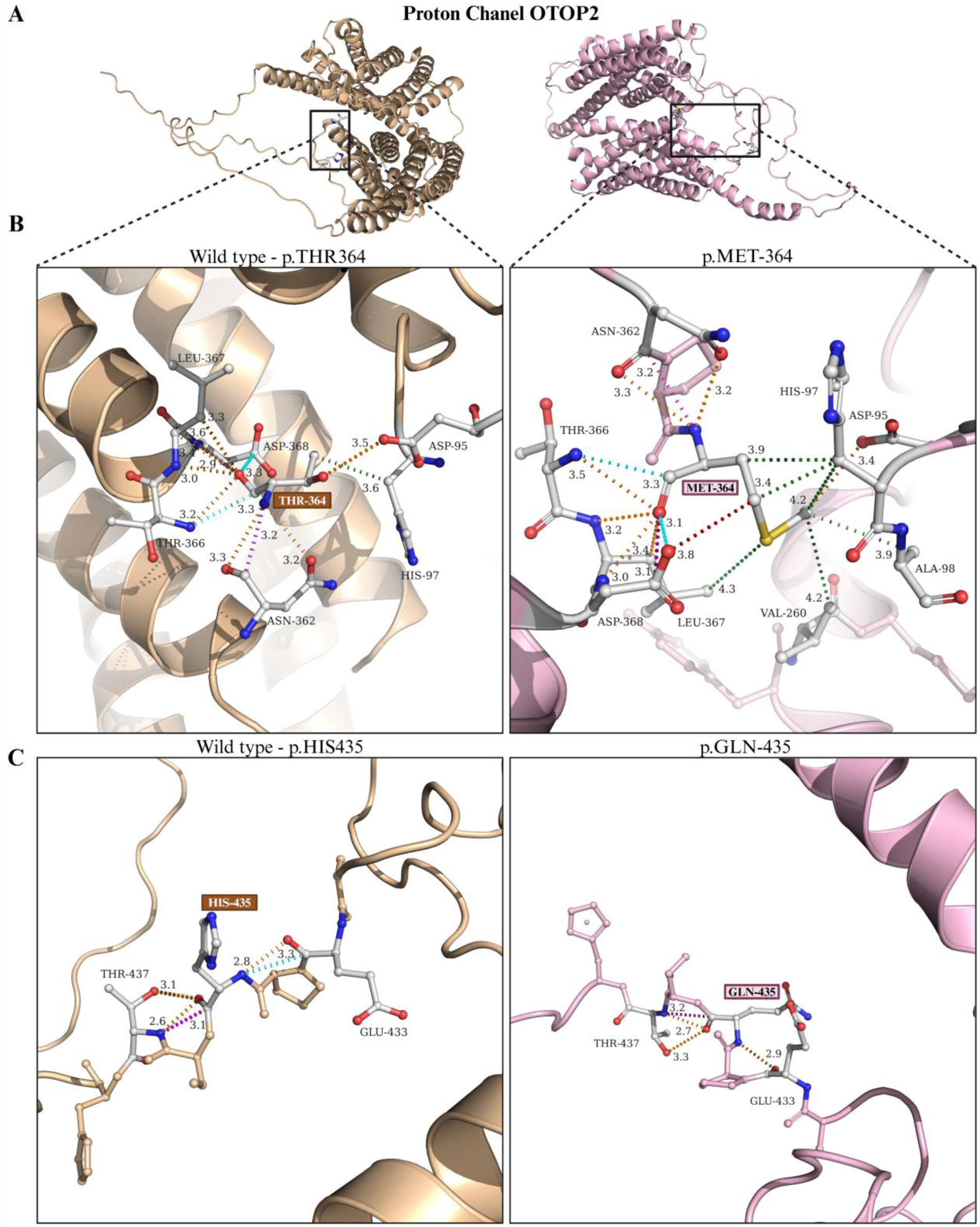
(A) Wild type (beige) and mutant (purple) proton channel OTOP2 protein models. (B) Atomic interactions between wild type Thr364 and mutant Met364. (C) Atomic interactions between wild type His435 and mutant Gln435. Mutated amino acids are highlighted.

Atomic interaction and stability analysis revealed that the p.Thr272Ile variant retained the same number of atomic interactions with neighbouring amino acids. In the wild-type protein, Thr-272 created a Van der Waal (VDW) interaction with Gly-274, as well as a polar contact and a clash with Ala-270. On the mutant protein, Ile-272 created a VDW interaction and apolar contacts with Ala-270 and a polar contact with Gly-274 (Figure 3B), leading to a less stable structure with a predicted Gibbs free energy (ΔG) change of -0.31 kcalmol^-1^. On the other hand, p.Thr364Met and p.His435Gln created a more stable structure with a ΔG change of 0.8 kcalmol^-1^ and 0.54 kcalmol^-1^, respectively. p.Thr364Met has a complex panorama of atomic interactions, with multiple bonds formed between Met-364 and neighbouring amino acids (Table S4). For p.His435Gln, the atomic interactions are largely preserved, except for the loss of a VDW interaction between Gln-435 and Glu-433 (Figure 4B-C).

## DISCUSSION

This study replicates the previously reported *MYO7A*-mediated digenic inheritance in MD and provides new findings supporting *OTOP2* as a novel MD candidate gene, showing a significant burden with missense variants in SK-MD individuals. Although the *OTOP2* variants reported in SK individuals are different from one previously reported in a sporadic case from Spain [10], the potential genetic contribution of *OTOP2* to MD pathogenesis deserves further investigations, since this biallelic inheritance pattern has been also observed in *OTOG*, the most common gene found in familial MD with European ancestry [7].

*OTOP2* encodes proton channel OTOP2 (also known as otopetrin-2), a proton-selective channel primarily expressed in the supporting cells of the mouse and human utricle ([36]; gEAR: https://umgear.org). Its homolog, *OTOP1*, is essential for otoconia formation and vestibular function in mice [37], suggesting a shared role for *OTOP2*. Notably, *OTOP2* variants are previously implicated in non-syndromic vestibular dysfunction [10] and Usher syndrome through its genomic proximity and CTCF-mediated regulatory interaction with *USH1G* [38,39]. While direct evidence of *OTOP2* clinical significance in MD pathogenicity remains to be established in larger samples, its expression in mammalian utricle and its emerging associations with MD-related phenotypes suggest a potential contribution to MD.

Although a burden of missense variants was observed in *OTOP2*, they are located in high-density regions (low constrained regions) within the EAS population (Figure 2A), supporting that these *OTOP2* variants may be tolerated by the protein. Among the identified variants, p.Thr272Ile is predicted to destabilize protein structure and potentially impair proton channel function. The p.His435Gln variant, located approximately 210 bp from a canonical splice site, possibly introduces a novel acceptor site. Given their potential impacts on protein structure and function, these variants could contribute to MD susceptibility and warrant further functional validation.

Of the remaining genes showing significant burden, *CCNJL*, *CLU*, and *FBP2* are primarily expressed in hair cells, while *CDON* and *BAHCC1* are mainly found in the supporting cells of cochlear and vestibular sensory epithelia during mouse embryo and postnatal development ([40–42]. These genes are involved in key cellular processes, such as cell cycle regulation (*CCNJL*), chromatin remodelling (*BAHCC1*), energy metabolism (*FBP2*), homeostasis (*CLU*), and Hedgehog signalling pathways (*CDON*), suggesting potential roles in inner ear development, despite limited evidence of their association with MD pathogenesis [40,41]. Besides, knockout models of genes showing significant burden, except *TMEM131L*, do not exhibit any hearing or vestibular phenotypes, as reported in International Mouse Phenotype Consortium (IMPC: https://mousephenotype.org). *TMEM131L* knockout in mice significantly impacts vestibular and hearing functions and is also predicted to be associated with SNHL in the IMPC mouse database. This gene is highly expressed in vestibular dark cells, which are involved in endolymph production – a key component of MD pathology [43–45]. Though its pathogenic role remains unclear, *TMEM131L* might share functional features with other MD-associated transmembrane proteins, which are associated with hearing loss and MD [46–48].

Two SK-MD individuals carry digenic variants in MD-linked genes, including *MYO7A*, *CDH23*, and *ADGRV1*, which are involved in stereocilia links and mechanoelectrical transduction complex of cochlear hair cells [49]. Notably, one *MYO7A* variant (chr11:77214688:G>A) matches a previously reported Spanish familial MD mutation [5,6]. The variants chr11:77172799:T>C in *MYO7A* (ClinVar ID: 438172) and chr10:71778252:G>A in *CDH23* (ClinVar ID: 197422) have been expert-curated, linking to hearing loss. These findings support the digenic inheritance pattern in MD.

Intriguingly, one rare variant in *OTOP2*, predicted to destabilise protein structure, co-occurs with digenic mutations in *MYO7A* in the same SK-MD individual, supporting a polygenic contribution. However, the cMAF of digenic mutations is higher than the prevalence of MD (1/10,000), although both variants have high CADD scores (CADD > 20). More SK-MD sequencing datasets and segregation studies are needed to validate the pathogenicity of *MYO7A*, and determine which mutation plays the primary pathogenic role.

Two SK-MD individuals, P1 and P2, were classified as sporadic cases with SNHL and episodic vertigo, lacking familial MD evidence as their family members only exhibit SNHL. While segregation data are currently limited, the presence of compound heterozygous and digenic missense variants supports its potential contribution to familial MD.

This study has several limitations. The relatively small SK-MD cohort limits statistical power for the single-variant association and gene burden tests. Only a small overlap (0.8%) of rare variants is observed with the previously reported variants in the Spanish cohort ([50]; Supplementary Fig. S5), with 2.6 times higher in the number of rare variants detected. However, most SK-specific variants have insignificant functional impact and most genes harbouring missense or LoF variants are shared between two population cohorts. A likely explanation for this divergence is the relatively small sample size of SK-MD cohort compared to Spanish cohort, which leads to reduced power of rare variant detection, but also the lower number of individuals in the EAS population compared to the NFE reported in gnomAD. Therefore, more EAS MD individuals should be collected to support findings of the study. Besides, the absence of parental genotypes limits segregation analyses and conclusions on inheritance patterns. This study also focuses on exonic variants and do not assess non-coding or regulatory regions.

To conclude, the presence of rare compound and digenic variants in known and novel genes, especially *OTOP2*, supports the hypothesis of a multiallelic and polygenic inheritance model in MD genetics. Functional modelling and gene expression analyses suggest a potential mechanistic link between disrupted proton channel and MD susceptibility, meriting its consideration as a novel MD gene. Further sequencing and segregation studies in the EAS population are needed to cover the existing gap and to refine our understanding of MD genetic structure, particularly in the EAS population.

## Data Availability

The data that support the findings of this study are openly available in ClinVar at https://www.ncbi.nlm.nih.gov/clinvar/, under the submission number XXXX.

## Code Availability

The code to analyse the data generated in this study is available at https://github.com/TamPham271299/germline-VC.

## Acknowledgement

We would like to acknowledge the patients that donated blood for this study.

## Contributions

M.T.P – Design of methodology. Curation, analysis, and interpretation of data. Writing – original draft. Writing – review & editing.

P.C.G – Analysis and interpretation of data. Writing – original draft. Writing – review & editing.

S.H.J – Conception and acquisition of data.

H.Y.G – Conception and acquisition of data.

J.J – Conception and acquisition of data. Reviewing the content. Final approval of the version to be published.

J.Y.C – Conception and acquisition of data.

S.H.K – Conception and acquisition of data. Reviewing the content. Final approval of the version to be published.

J.A.L.E – Conception of data. Supervision. Reviewing the content. Funding acquisition.

All authors have read and approved the final version of the manuscript.

## Ethical Approval

This study was approved by the Human Ethics Committee at The University of Sydney (2023/HE000199) and the human research institutional review boards at the Yonsei University College of Medicine, Seoul, Korea (IRB #4-2015-0659) and was planned according to the standards of the Declaration of Helsinki. Patient information and written consent were obtained from all participants.

