## Supplementary Material for "Rare Missense Variants in *MYO7A* and *OTOP2* Genes in a South Korean Meniere Disease Cohort"

**Table S1:** Gene burden showing enrichment of missense variants in SK patients with MD. Genes are ordered by descending adjusted P-values of Fisher’s exact test of SK-MD vs. EAS gnomAD.

| **Gene** | **No. of variants** | **No. of individuals** | **Variants** | **AF** | | | **gnomAD (EAS)** | | | **gnomAD (Global)** | | |
| --- | --- | --- | --- | --- | --- | --- | --- | --- | --- | --- | --- | --- |
|  |  |  |  | **SK-MD** | **gnomAD (EAS)** | **gnomAD (Global)** | **OR (95% CI)** | **adj. P-value** | **EF** | **OR (95% CI)** | **adj. P-Value** | **EF** |
| *CCNJL* | 3 | 3 | chr5:160280546:C>T | 0.031 | 1.01x10^-4^ | 3.77 x10^-5^ | 774.41  (117.33-3884.09) | 1.30x10^-4^ | 1 | 1315.88  (258.42-4276.05) | 1.06x10^-5^ | 1 |
|  |  |  | chr5:160280587:C>T | 0.031 | 0 | 2.40 x10^-5^ |  |  |  |  |  |  |
|  |  |  | chr5:160280627:G>A | 0.031 | 2.52 x10^-5^ | 1.30 x10^-5^ |  |  |  |  |  |  |
| *BAHCC1* | 4 | 4 | chr17:81443334:G>A | 0.031 | 1.66 x10^-4^ | 9.57 x10^-6^ | 71.61  (18.6-196.47) | 2.26x10^-3^ | 0.99 | 3.41  (0.91-8.95) | n.s. | 0.71 |
|  |  |  | chr17:81443502:G>A | 0.031 | 1.46 x10^-3^ | 1.13 x10^-4^ |  |  |  |  |  |  |
|  |  |  | chr17:81460539:C>T | 0.031 | 0 | 1.67 x10^-5^ |  |  |  |  |  |  |
|  |  |  | chr17:81463711:G>A | 0.031 | 2.77 x10^-4^ | 3.58 x10^-2^ |  |  |  |  |  |  |
| *IGFN1* | 6 | 5 | chr1:201208192:T>A | 0.031 | 8.12 x10^-4^ | 2.31 x10^-5^ | 19.7  (7.09-44.14) | 4.58x10^-3^ | 0.95 | 178.6  (64.48-396.62) | 1.01x10^-8^ | 0.99 |
|  |  |  | chr1:201211942:C>T | 0.031 | 3.67 x10^-3^ | 7.56 x10^-4^ |  |  |  |  |  |  |
|  |  |  | chr1:201212092:C>T | 0.031 | 2.52 x10^-3^ | 1.05 x10^-4^ |  |  |  |  |  |  |
|  |  |  | chr1:201213327:C>G | 0.031 | 1.43 x10^-3^ | 4.24 x10^-5^ |  |  |  |  |  |  |
|  |  |  | chr1:201215148:G>A | 0.031 | 1.49 x10^-3^ | 5.82 x10^-5^ |  |  |  |  |  |  |
|  |  |  | chr1:201225875:C>T | 0.031 | 1.01 x10^-4^ | 1.18 x10^-4^ |  |  |  |  |  |  |
| *CDON* | 2 | 1 | chr11:125961901:G>A | 0.031 | 0 | 6.84 x10^-7^ | 2436.47  (130.24-4.50x10^15^) | 8.80x10^-3^ | 1 | 16383.64  (2871.61-4.50x10^15^) | 1.30x10^-5^ | 1 |
|  |  |  | chr11:126005932:G>A | 0.031 | 2.52 x10^-5^ | 6.84 x10^-7^ |  |  |  |  |  |  |
| *CLU* | 2 | 2 | chr8:27599971:G>A | 0.031 | 2.52 x10^-5^ | 1.37 x10^-5^ | 1256.98  (90.63-16384) | 1.76x10^-2^ | 1 | 3444.41  (424.88-16384) | 7.61x10^-4^ | 1 |
|  |  |  | chr8:27605073:C>T | 0.031 | 2.52 x10^-5^ | 3.42 x10^-6^ |  |  |  |  |  |  |
| *UROC1* | 2 | 2 | chr3:126498125:G>A | 0.031 | 0 | 7.52 x10^-6^ | 1256.98  (90.63-16384) | 1.76x10^-2^ | 1 | 2145.48  (257.38-8192) | 2.05x10^-3^ | 1 |
|  |  |  | chr3:126505766:C>T | 0.031 | 5.04 x10^-5^ | 2.12 x10^-5^ |  |  |  |  |  |  |
| *GGT7* | 2 | 2 | chr20:34852443:G>A | 0.031 | 0 | 2.05 x10^-5^ | 1253.1  (90.31-16384) | 1.77x10^-2^ | 1 | 3178.03  (334.41-16384) | 1.22x10^-3^ | 1 |
|  |  |  | chr20:34863474:C>G | 0.031 | 5.07 x10^-5^ | 1.37 x10^-6^ |  |  |  |  |  |  |
| *MTCL2* | 3 | 3 | chr20:36794141:T>C | 0.031 | 5.32 x10^-4^ | 1.36 x10^-5^ | 112.23  (21.64-370.85) | 1.81x10^-2^ | 0.99 | 4468.26  (819.51-16384) | 3.50x10^-7^ | 1 |
|  |  |  | chr20:36803061:T>C | 0.031 | 7.59 x10^-5^ | 2.06 x10^-6^ |  |  |  |  |  |  |
|  |  |  | chr20:36808587:T>C | 0.031 | 2.77 x10^-4^ | 7.54 x10^-6^ |  |  |  |  |  |  |
| *OTOP2* | 3 | 2 | chr17:74930450:C>T | 0.031 | 4.79 x10^-4^ | 1.57 x10^-5^ | 100.88  (19.6-326.87) | 2.43x10^-2^ | 0.99 | 1057  (225.35-3834.66) | 1.63x10^-5^ | 1 |
|  |  |  | chr17:74930726:C>T | 0.031 | 7.56 x10^-5^ | 3.28 x10^-5^ |  |  |  |  |  |  |
|  |  |  | chr17:74930940:C>G | 0.031 | 4.03 x10^-4^ | 3.76 x10^-5^ |  |  |  |  |  |  |
| *TMEM131L* | 4 | 4 | chr4:153585489:T>G | 0.031 | 7.31 x10^-4^ | 2.74 x10^-5^ | 37.92  (10.02-102.04) | 2.53x10^-2^ | 0.97 | 1305.08  (333.24-3815.33) | 2.20x10^-8^ | 1 |
|  |  |  | chr4:153603428:A>T | 0.031 | 1.89 x10^-3^ | 5.20 x10^-5^ |  |  |  |  |  |  |
|  |  |  | chr4:153604177:T>G | 0.031 | 4.03 x10^-4^ | 1.23 x10^-5^ |  |  |  |  |  |  |
|  |  |  | chr4:153635501:C>G | 0.031 | 3.78 x10^-4^ | 1.03 x10^-5^ |  |  |  |  |  |  |
| *ADGRG3* | 2 | 2 | chr16:57676216:G>A | 0.031 | 5.04 x10^-5^ | 4.10 x10^-6^ | 852.65  (69.74-8192) | 2.93x10^-2^ | 1 | 1888.94  (231.13-8192) | 2.55x10^-3^ | 1 |
|  |  |  | chr16:57680545:G>A | 0.031 | 2.52 x10^-5^ | 2.80 x10^-5^ |  |  |  |  |  |  |
| *TENM2* | 3 | 3 | chr5:168218284:C>G | 0.031 | 9.32 x10^-4^ | 2.53 x10^-5^ | 84.97  (16.6-270.7) | 3.96x10^-2^ | 0.99 | 362.15  (73.79-1098.68) | 4.55x10^-4^ | 1 |
|  |  |  | chr5:168218630:T>A | 0.031 | 2.52 x10^-5^ | 6.84 x10^-7^ |  |  |  |  |  |  |
|  |  |  | chr5:168262250:G>A | 0.031 | 1.78 x10^-4^ | 2.39 x10^-4^ |  |  |  |  |  |  |
| *FBP2* | 2 | 2 | chr9:94559071:G>A | 0.031 | 0 | 3.35 x10^-5^ | 631.56  (56.7-4026.34) | 4.40x10^-2^ | 1 | 1743.61  (202.78-8192) | 3.34x10^-3^ | 1 |
|  |  |  | chr9:94587341:T>C | 0.031 | 1.01 x10^-4^ | 3.42 x10^-6^ |  |  |  |  |  |  |
| *GPR180* | 2 | 2 | chr13:94619248:A>G | 0.031 | 5.04 x10^-5^ | 1.37 x10^-6^ | 631.24  (56.67-4025.14) | 4.40x10^-2^ | 1 | 10922.43  (1463.18-4.50x10^15^) | 6.08x10^-5^ | 1 |
|  |  |  | chr13:94623284:A>G | 0.031 | 5.04 x10^-5^ | 2.74 x10^-6^ |  |  |  |  |  |  |

EAS: East Asian; adj. P-value: adjusted P-value after Bonferroni correction; OR (95% CI): Odd Ratio (95% Confidence Interval); EF: Etiological Fraction; n.s.: non-significant (adj. P-value < 0.05)

**Table S2:** Splicing prediction by *SpliceAI* and *Pangolin* for OTOP2 variants.

| **Variant** | **Prediction Tool** | **Δ  type** | **Δ  score** | **Position** |
| --- | --- | --- | --- | --- |
| chr17:74930726C>T | SpliceAI | Acceptor Loss | 0 |  |
|  |  | Donor Loss | 0 | 41 bp |
|  |  | Acceptor Gain | 0 |  |
|  |  | Donor Gain | 0 |  |
|  | Pangolin | Splice Loss | 0 | 41 bp |
|  |  | Splice Gain | 0 | -299 bp |
| chr17:74930450C>T | SpliceAI | Acceptor Loss | 0 | -194 bp |
|  |  | Donor Loss | 0.01 | -42 bp |
|  |  | Acceptor Gain | 0 | 0 |
|  |  | Donor Gain | 0 | 0 |
|  | Pangolin | Splice Loss | 0.01 | -194 bp |
|  |  | Splice Gain | 0 | -42 bp |
| chr17:74930940C>G | SpliceAI | Acceptor Loss | 0 |  |
|  |  | Donor Loss | 0 | -173 bp |
|  |  | Acceptor Gain | 0 |  |
|  |  | Donor Gain | 0 | 207 bp |
|  | Pangolin | Splice Loss | 0 | 3 bp |
|  |  | Splice Gain | 0 | 213 bp |

**Table S3:** Creation and deletion of Exonic Splicing Enhancers (ESE) and Exonic Splicing Silencers (ESS) regulatory elements in *OTOP2* variants.

| **Variant** | **Regulatory Element** | **Name** | **Position** | **Sequence** | **Status** |
| --- | --- | --- | --- | --- | --- |
| chr17:74930450C>T | ESS | Sironi motif3 | chr17:74930443 | GCCTCCAC | Site Broken |
|  | ESE | RESCUE ESE | chr17:74930445 | CTCCAT | Site Created |
|  | ESE | ESE_SRp55 | chr17:74930446 | TCCATC | Site Created |
|  | ESE | EIE | chr17:74930447 | CCACCC | Site Broken |
|  | ESE | ESE_SRp40 | chr17:74930447 | CCACCCC | Site Broken |
|  | ESS | Sironi_motif3 | chr17:74930447 | CCACCCCT | Site Broken |
|  | ESE | EIE | chr17:74930448 | CACCCC | Site Broken |
|  | ESE | ESE_ASF | chr17:74930448 | CACCCCT | Site Broken |
|  | ESE | ESE_ASFB | chr17:74930448 | CACCCCT | Site Broken |
|  | ESS | Sironi_motif3 | chr17:74930448 | CATCCCTG | Site Created |
|  | ESE | ESE_ASFB | chr17:74930450 | CCCCTGG | Site Broken |
| chr17:74930726C>T | ESE | EIE | chr17:74930721 | CCCCAC | Site Broken |
|  | ESE | ESE_SRp40 | chr17:74930721 | CCCCACG | Site Broken |
|  | ESE | ESE_ASF | chr17:74930722 | CCCACGC | Site Broken |
|  | ESE | ESE_ASFB | chr17:74930722 | CCCACGC | Site Broken |
|  | ESE | ESE_SRp40 | chr17:74930723 | CCACGCG | Site Broken |
|  | ESE | ESE_ASFB | chr17:74930724 | CACGCGC | Site Broken |
|  | ESS | Sironi_motif3 | chr17:74930725 | ACGCGCAC | Site Broken |
|  | ESE | ESE_SRp55 | chr17:74930726 | TGCGCA | Site Created |
| chr17:74930940C>G | ESE | ESE_SC35 | chr17:74930933 | AGCCTCAG | Site Created |
|  | ESE | EIE | chr17:74930935 | CCTCAC | Site Broken |
|  | ESE | EIE | chr17:74930936 | CTCAGA | Site Created |
|  | ESE | ESE_ASF | chr17:74930936 | CTCACAG | Site Broken |
|  | ESE | ESE_SRp40 | chr17:74930937 | TCACAGT | Site Broken |
|  | ESS | Sironi_motif1 | chr17:74930937 | TCAGAGTA | Site Created |
|  | ESS | ESS_hnRNPA1 | chr17:74930938 | CAGAGT | Site Created |
|  | ESE | ESE_9G8 | chr17:74930940 | GAGTAC | Site Created |

**Table S4:** Atomic Interactions in *OTOP2* p.T364M and Neighbouring Amino Acids

| **Protein Variant** | **Amino Acid - Amino Acid** | **Atom - Atom** | **Type of Interaction** | **Distance (Å)** |
| --- | --- | --- | --- | --- |
| **Wild type** | Thr364-Asp95 | OG1 - OD2 | Polar Contact | 3.5 |
|  | Thr364-His97 | CG2 - CB | Hydrophobic Interaction | 3.6 |
|  | Thr364-Asn362 | N-C | Clash | 3.2 |
|  | Thr364-Asn362 | N-O | Polar Contact | 3.3 |
|  | Thr364-Asn362 | N-OD1 | Polar Contact | 3.2 |
|  | Thr364-Thr366 | C-N | Van der Waals Interaction | 3.3 |
|  | Thr364-Thr366 | O-N | Polar Contact | 3.2 |
|  | Thr364-Leu367 | O-N | Polar Contact | 3.0 |
|  | Thr364-Leu367 | O-CA | Polar Contact | 3.4 |
|  | Thr364-Leu367 | O-CB | Polar Contact | 3.3 |
|  | Thr364-Leu367 | O-C | Carbonyl Interaction | 3.6 |
|  | Thr364-Asp368 | O-N | Polar Contact | 2.9 |
|  | Thr364-Asp368 | O-CG | Van der Waals Interaction | 3.3 |
| **Mutant** | Met364-Asp95 | CE-OD2 | Polar Contact | 3.4 |
|  | Met364-His97 | CE-CB | Hydrophobic Interaction | 4.2 |
|  | Met364-His97 | CG-CB | Hydrophobic Interaction | 3.4 |
|  | Met364-His97 | CB-CB | Hydrophobic Interaction | 3.9 |
|  | Met364-Ala98 | CE-CB | Hydrophobic Interaction | 3.9 |
|  | Met364-Val260 | CE-CG1 | Hydrophobic Interaction | 4.2 |
|  | Met365-Asn362 | N-OD1 | Polar Contact | 3.2 |
|  | Met365-Asn362 | N-C | Clash | 3.2 |
|  | Met365-Asn362 | N-O | Polar Contact | 3.3 |
|  | Met354-Thr366 | C-N | Van der Waals Interaction | 3.3 |
|  | Met354-Thr366 | O-N | Polar Contact | 3.5 |
|  | Met365-Leu367 | SD-CD2 | Hydrophobic Interaction | 4.3 |
|  | Met365-Leu367 | O-CB | Polar Contact | 3.4 |
|  | Met365-Leu367 | O-N | Polar Contact | 3.2 |
|  | Met365-Thr368 | O-N | Polar Contact | 3.0 |
|  | Met365-Thr368 | O-CG | Clash | 3.1 |
|  | Met365-Thr368 | O-OD2 | Van der Waals Interaction | 3.1 |
|  | Met365-Thr368 | CG-OD2 | Hydrogen Bond | 3.8 |

**Supplementary Figure S1:** Bioinformatic analysis pipeline

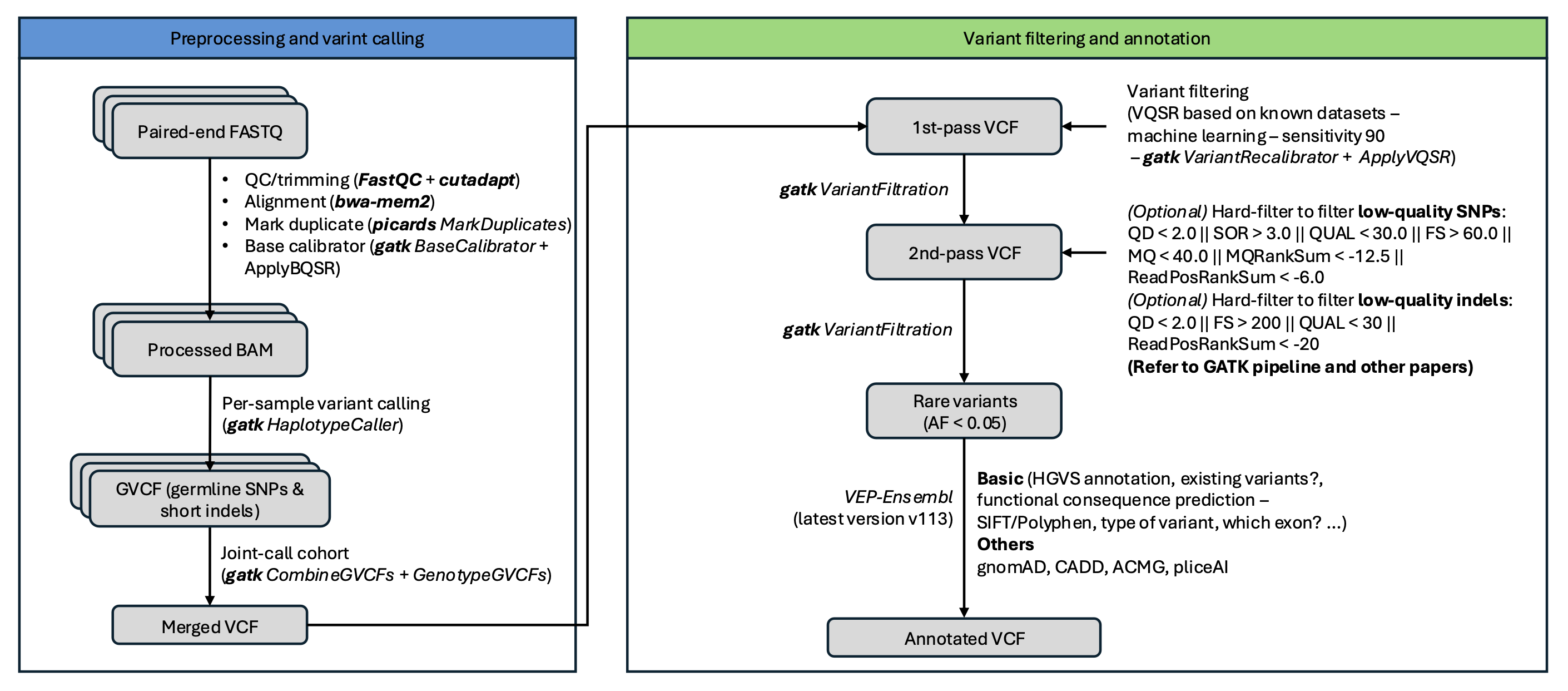

**Supplementary Figure S2:** Pathogenicity Heatmap for amino acid change p.T272I in *OTOP2* canonical transcript, indicating a low pathogenic score site of 0.0.88.

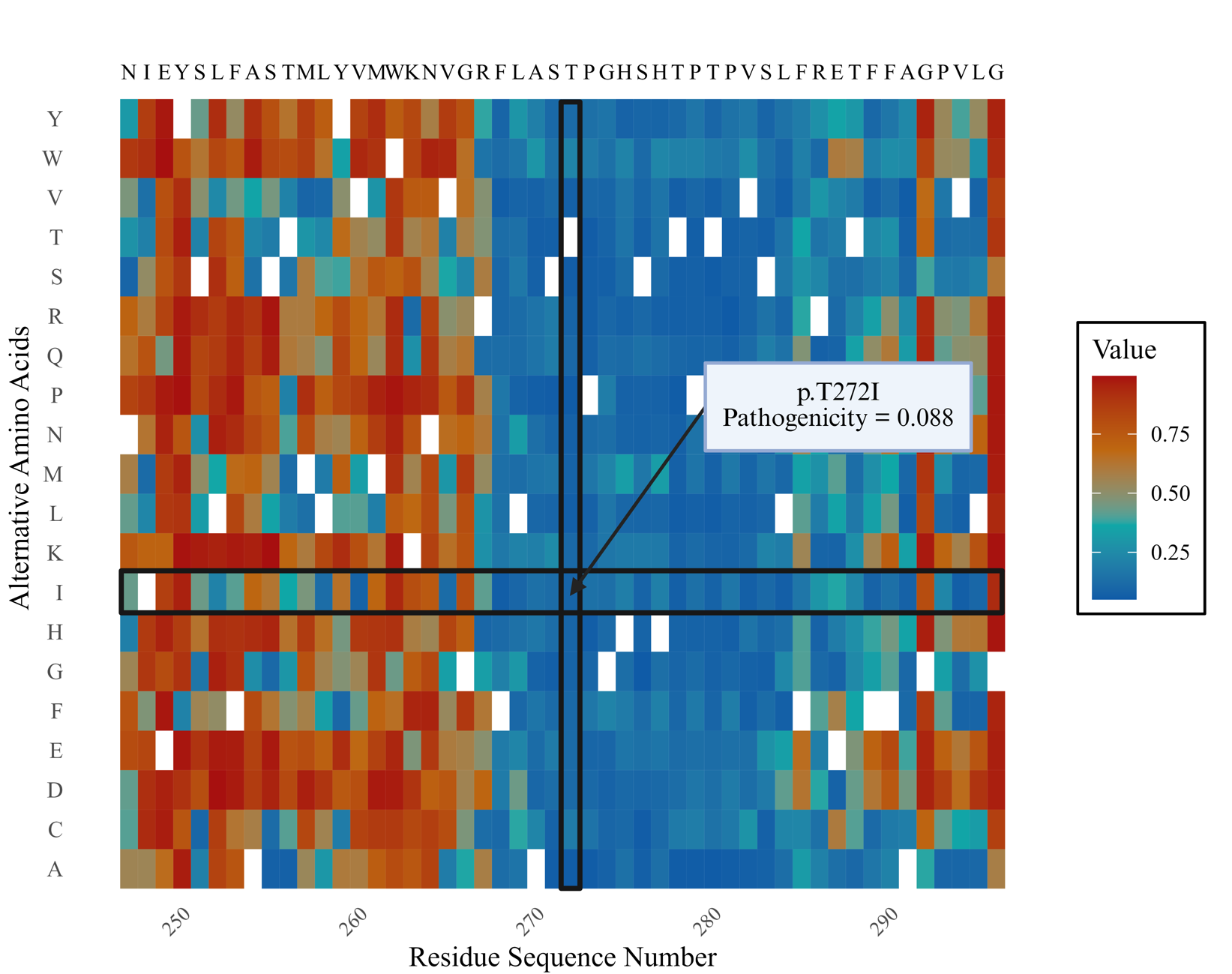

**Supplementary Figure S3:** Pathogenicity Heatmap for amino acid change p.T364M in *OTOP2* canonical transcript, indicating a low pathogenic score site of 0.141.

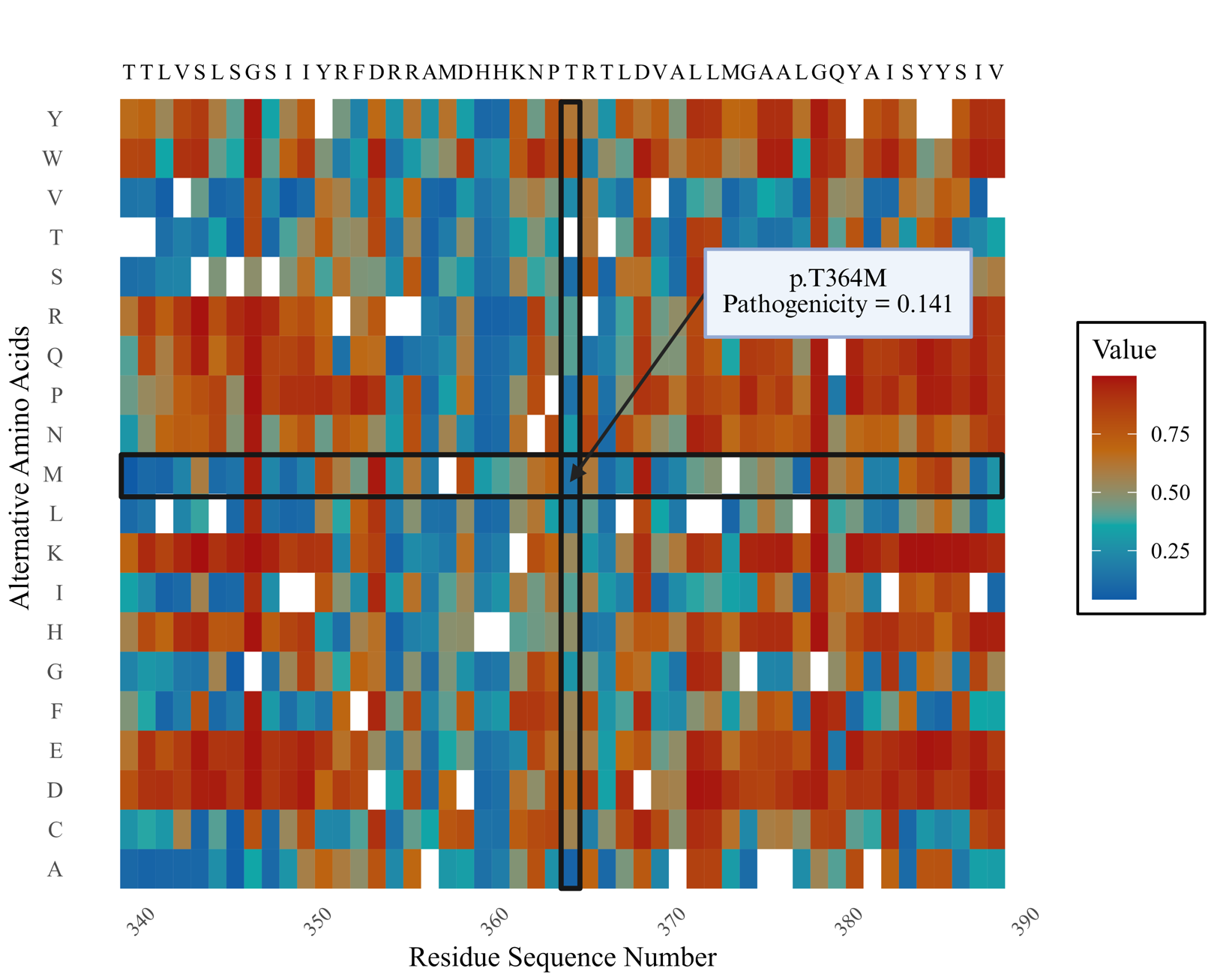

**Supplementary Figure S4:** Pathogenicity Heatmap for amino acid change p.H435 in *OTOP2* canonical transcript, indicating a low pathogenic score site of 0.079.

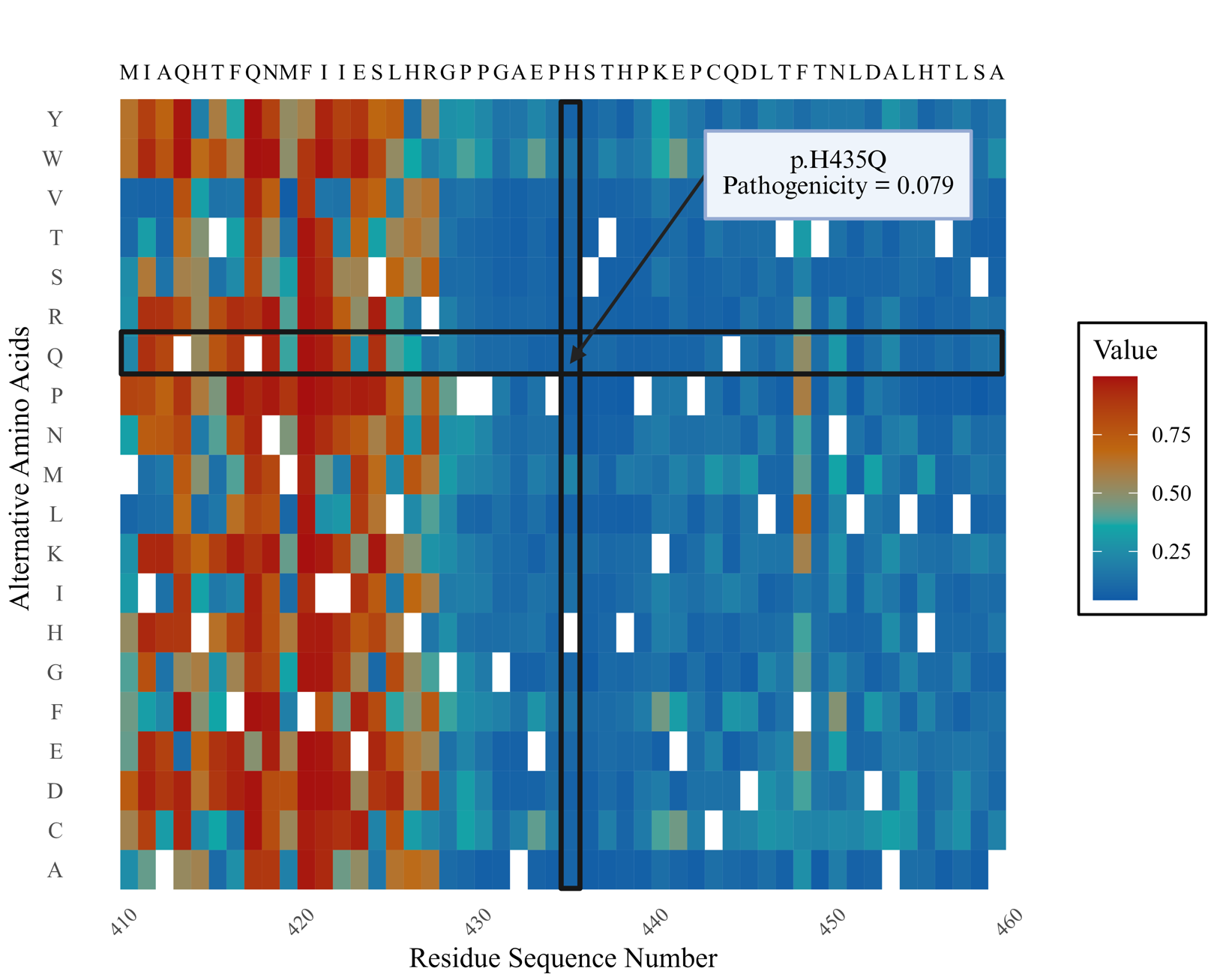

**Supplementary Figure S5:** Overlaps of (A) rare variants in five functional categories and (B) genes with only rare missense and LoF between the SK and Spain MD cohort.

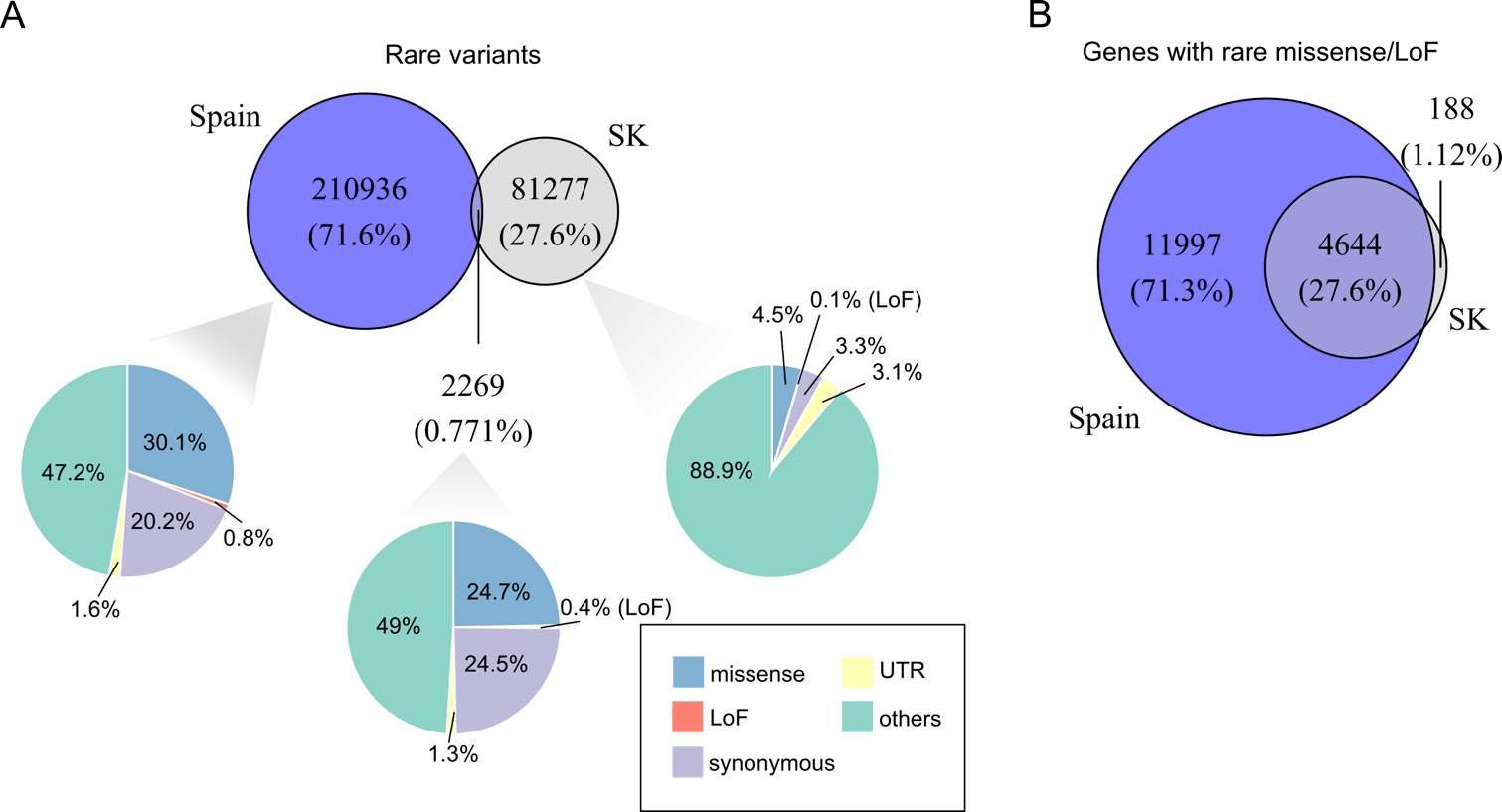
